# Accounting for underreporting in mathematical modelling of transmission and control of COVID-19 in Iran

**DOI:** 10.1101/2020.05.02.20087270

**Authors:** Meead Saberi, Homayoun Hamedmoghadam, Kaveh Madani, Helen M. Dolk, Andrei S. Morgan, Joan K. Morris, Kaveh Khoshnood, Babak Khoshnood, on behalf of the Iran COVID-19 working group

## Abstract

**Background:** Iran has been the hardest hit country by the outbreak of SARS-CoV-2 in the Middle East with 74,877 confirmed cases and 4,683 deaths as of 15 April 2020. With a relatively high case fatality ratio and limited testing capacity, the number of confirmed cases reported is suspected to suffer from significant under-reporting. Therefore, understanding the transmission dynamics of COVID-19 and assessing the effectiveness of the interventions that have taken place in Iran while accounting for the uncertain level of underreporting is of critical importance. We use a mathematical epidemic model utilizing official confirmed data and estimates of underreporting to understand how transmission in Iran has been changing between February and April 2020.

**Methods:** We developed a compartmental transmission model to estimate the effective reproduction number and its fluctuations since the beginning of the outbreak in Iran. We associate the variations in the effective reproduction number with a timeline of interventions and national events. The estimation method also accounts for the underreporting due to low case ascertainment by estimating the percentage of symptomatic cases using delay-adjusted case fatality ratio based on the distribution of the delay from hospitalization-to-death.

**Findings:** Our estimates of the effective reproduction number ranged from 0.66 to 1.73 between February and April 2020, with a median of 1.16. We estimate a reduction in the effective reproduction number during this period, from 1.73 (95% CI 1.60 – 1.87) on 1 March 2020 to 0.69 (95% CI 0.68-0.70) on 15 April 2020, due to various non-pharmaceutical interventions including school closures, a ban on public gatherings including sports and religious events, and full or partial closure of non-essential businesses. Based on these estimates and given that a near complete containment is no longer feasible, it is likely that the outbreak may continue until the end of the 2020 if the current level of physical distancing and interventions continue and no effective vaccination or therapeutic are developed and made widely available.

**Interpretation:** The series of non-pharmaceutical interventions and the public compliance that took place in Iran are found to be effective in slowing down the speed of the spread of COVID-19 within the studied time period. However, we argue that if the impact of underreporting is overlooked, the estimated transmission and control dynamics could mislead the public health decisions, policy makers, and general public especially in the earlier stages of the outbreak.

**Funding:** Nil.

## RESEARCH IN CONTEXT

### Evidence before this study

Since the outbreak of SARS-CoV-2 in late 2019, several studies have attempted to understand its transmission and control dynamics. The majority of the existing studies reported the dynamics and initial estimates of the effective reproduction number from China followed by a few other studies using data from other countries including Italy, Spain, South Korea, Germany, France and Iran among others. However, none of the previous work has taken into account the impact of possible underreporting of cases in estimation of the effective reproduction number. Also, no other study reported the time-dependent association between the interventions and variations in the effective reproduction number for Iran as the hardest hit country in the Middle East.

### Added value of this study

We use a mathematical model to estimate the transmission and control dynamics of COVID-19 in Iran, taking into account the significant underreporting of cases. We estimated the time-dependent effective reproduction number in association with a timeline of events and interventions that took place. We showed that if underreporting is overlooked, the estimated dynamics could mislead the public health decisions and general public. The impact of control measures on the effective reproduction number could also significantly be overestimated unless under-reporting is taken into account.

### Implications of all the available evidence

The estimation of transmission and control dynamics of COVID-19 in any country highly depends on the quality of the reported data. However, in the presence of high uncertainty in the number of confirmed cases, it is of crucial importance to take into account the impact of underreporting when interventions are to be introduced or lifted.

## INTRODUCTION

The outbreak of SARS-CoV-2 in Iran was first officially announced in February 2020, two months after the initial outbreak in Wuhan, China.^1^ Iran’s patient zero is believed to have been a merchant from Qom who had travel history to China.^2^ Despite the initial signs of a spread in Qom, the government declined to place the city under quarantine to contain the epidemic at an early stage for various technical, socio-economic, religious, and security reasons.^3^ The first local non-pharmaceutical interventions such as schools and universities closure were put in place a few days after the official acknowledgement of the first cases in Qom and Tehran.^4^ Since then, various public health control measures at the local and national levels were taken that are believed to have altered the course of the outbreak. See figure 1 for a spatial illustration of the spread throughout the country by province in the first week since the official announcement of the first case (appendix p 1). The relatively high case fatality ratio (CFR), defined as the total number of deaths over the total number of infected cases, in Iran’s official reports after the first week since the official declaration of the first case (16.8%) has raised questions on the true number of cases in the country.^5,6^ The testing protocol in Iran at the early stages of the outbreak was limited to hospital admissions of the patients with severe symptoms. While Iran has extended the COVID-19 diagnostic testing capacity later on to patients with milder symptoms, it is believed that under-ascertainment of cases still remains high.

**Figure 1:**
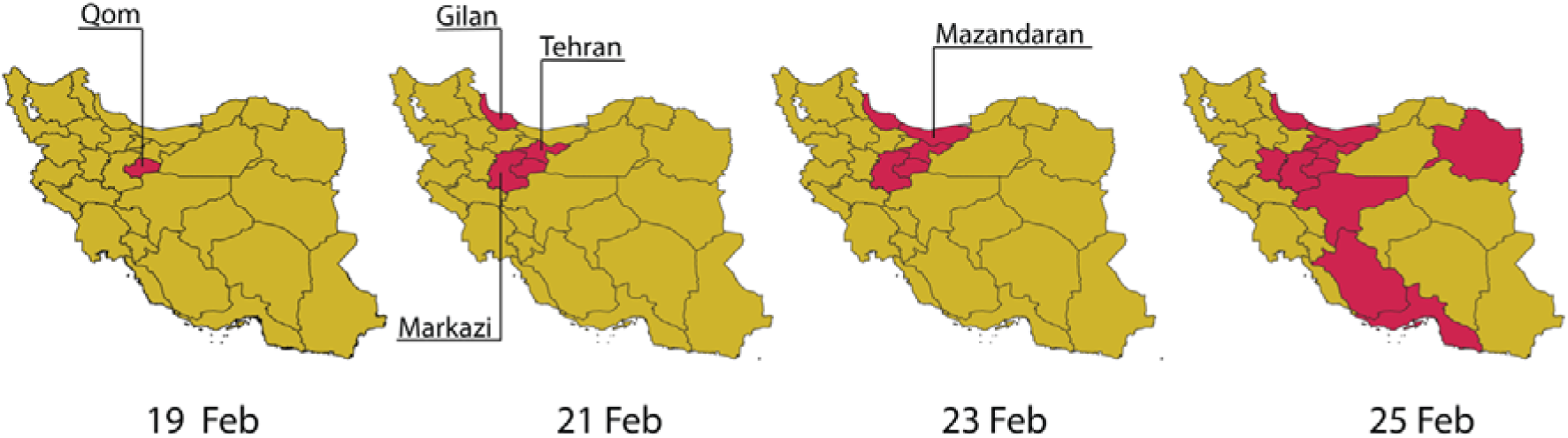
Spatial illustration of the spread of COVID-19 in Iran by province in the first week of the official outbreak announcement from 19 February to 25 February 2020.

This study aims to understand the transmission dynamics of COVID-19 in Iran and to assess the effectiveness of the control measures that were put in place over time through estimation of the effective reproduction number *R*(*t*) defined as the average number of susceptible persons infected by an infected person during its infectious period at a given time in the course of the epidemic. We assessed *R*(*t*) in relation to a timeline of national events and non-pharmaceutical interventions. In the absence of timely and reliable data, modelling can provide helpful answers, including the degree of plausible uncertainty in different estimates and the effectiveness of non-pharmaceutical interventions. By providing explicit and clear information about model assumptions and parameters, modelling can also foster scientific discussion of data gaps and what can be done to improve outbreak-related estimates by borrowing information available elsewhere. Finally, models can be developed and presented using both average estimates and measures of their uncertainty or, alternatively, as scenarios that can illustrate possible developments of the epidemic under various conditions.

## METHODS

### Data Sources

We use official time-series reports of the number of confirmed cases, recovered, and deaths from the World Health Organization (WHO)^1^ and Iran’s Ministry of Health and Medical Education^7^. The first confirmed case was reported on 19 February 2020 which is assumed as the beginning of the outbreak of COVID-19 in Iran.

### Mathematical Model

We describe the dynamics of spread using a variation of the susceptible-exposed-infected-recovered (SEIR) model, distinguishing between fatality and recovered cases combined with an estimate of the percentage of symptomatic cases using delay-adjusted CFR (appendix p 1). See figure 2. The model accounts for the time between exposure-to-onset of symptoms (or confirmation), also known as incubation period, assuming a gamma distribution with an average of 5.5 days and a standard deviation of 2.3 days.^8^ We also assume the time from symptoms onset-to-death and -to-recovery both follow a gamma distribution with an average of 22.3 days and a standard deviation of 9.4 days and an average of 22.2 days and a standard deviation of 10 days, respectively.^9^ The size of the initial susceptible population is assumed to be 80 million.

**Figure 2:**
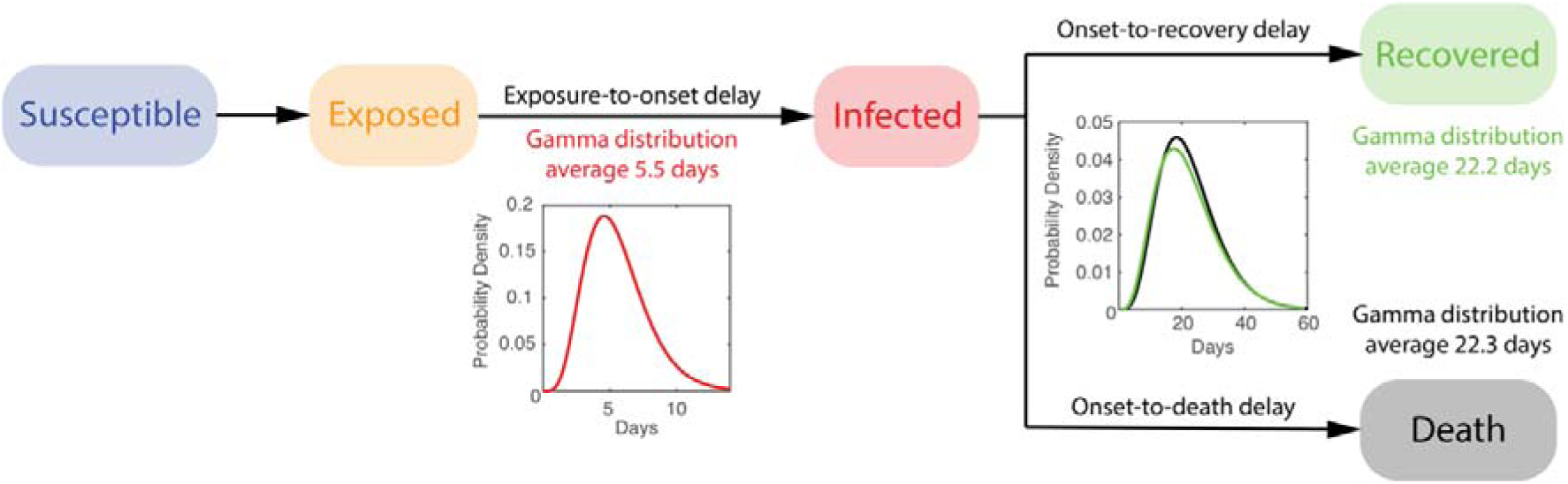
Schematic illustration of the formulated mathematical epidemic model.

### Parameter Estimation Method

To estimate the parameters of the developed SEIR model, we formulate an ordinary least squares (OLS) minimization.^10^ We use pattern search as a derivative-free global optimization algorithm^10^ to find the model parameters that minimizes the sum of the normalized root mean squared error (RMSE) of the number of infected *ε_i_* recovered *ε_r_* and removed cases *ε_f_*.

### Effective Reproduction Number

The basic reproduction number, *R*_0_, a fundamental measure in infectious disease epidemiology and public health, is defined as the average number of susceptible persons infected by an infected person during its infectious period in a fully susceptible population. *R*_0_ is known to fluctuate if the social contact rate in the population changes over time or space. Since *R*_0_ assumes an entirely susceptible population, several studies in the literature ^12–14^ suggest using the effective reproduction number *R_t_* when examining the effect of various interventions including vaccination, social distancing, and quarantine. *R_t_* is defined similarly to *R_0_* but is not limited to the assumption of a completely susceptible population. Here, we use empirical data from Iran to trace changes in *R_t_* over a rolling 7-day period since the beginning of the COVID-19 outbreak and describe its association with various interventions (e.g. school closures, social distancing, and bans on public gatherings) that took place by the public and government.

Various methods exist to estimate *R*_0_ (and *R_t_*).^15,16^ Here, we use the same framework described in figure 2 in which the parameters of the formulated SEIR model are inferred through an optimization problem. *R_t_* is calculated using a rolling time window of 7 days to capture the evolving trend of the spread over time due to various changes in the social network contact rate. The calculated *R_t_* may be overestimated during the early stage of an outbreak^17^ due to different reasons including the impact of imported cases and heterogeneity in subpopulations (e.g. older than 60 years old) with higher transmission rates.

### Accounting for Underreporting

We account for the under-reporting of the number of infected cases in the official confirmed data using delay-adjusted case fatality ratio (CFR) approach.^18^ This approach assumes that the time from hospitalization-to-death has a known statistical distribution and uses this distribution to estimate when the people who died from COVID-19 would have been reported as being infected. The case fatality ratio is the ratio of the numbers of deaths over the numbers of reported infections calculated at the time of reporting not the time of death. This is extremely important for rapidly evolving epidemics.

The method, however, does not account for underreporting in fatality cases. The distribution of the time from confirmation-to-death is assumed to follow the same distribution as of the time from hospitalization-to-death, following a lognormal distribution with a mean of 13 days and a standard deviation of 12.7 days.^19^ Here, we assume that the estimate for percentage of symptomatic COVID-19 cases reported for Iran follows a lognormal distribution (same distribution type as of the time from hospitalization-to-death) with a mean of 9.9% and a standard deviation of 4% based on the latest estimates in the literature.^18^ This is based on the assumption of a baseline CFR of 1.4%. We assume the underreporting level remains constant over time.

## RESULTS

We estimated that *R_t_* varied between 1.73 and 0.69 during March to April 2020 in Iran, with a median of 1.16. We estimated a reduction in *R_t_* from early-March to mid-April, from 1.35 (95% CI 1.31-1.39) on 8 March 2020 to 0.69 (95% 0.68-0.70) on 15 April 2020 due to control measures that took place. See figure 3(a) for a timeline of interventions and events and figure 3(b) for temporal variations of estimated *R_t_*.

**Figure 3:**
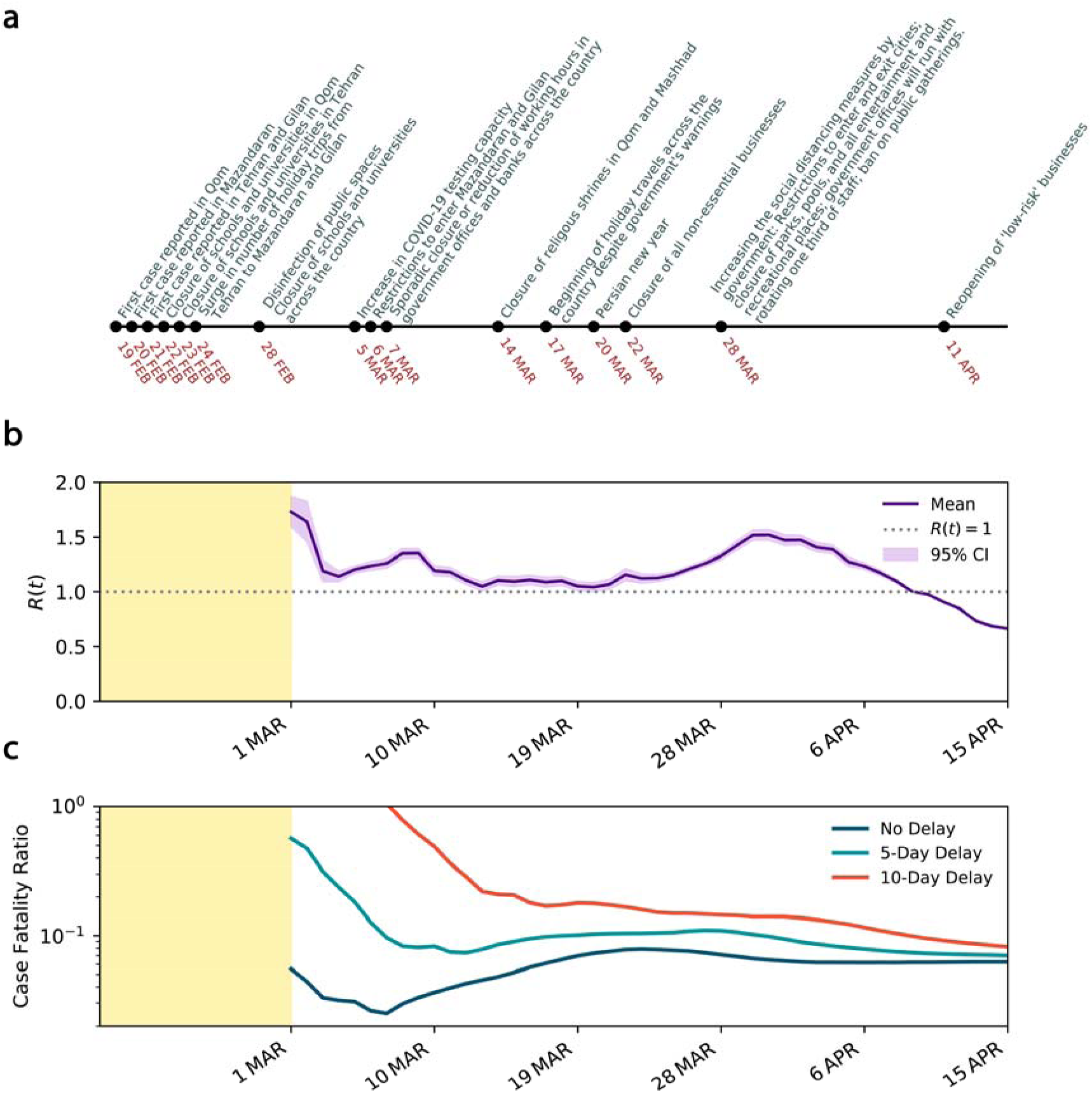
Temporal evolution of SARS-CoV-2 transmission characteristics in Iran. (a) Timeline of events and interventions that took place since the first case was reported on 19 February 2020. (b) Estimated effective reproduction number *R*(*t*) with 95% CI over a 7-day rolling window when underreporting is taken into account. (c) Estimated CFR with and without delay adjustment over the same time period in a semi-log scale.

The CFR on 25 February 2020, before adjusting for the time from diagnosis-to-death, was 16.84%. With more data emerging after the second week, the CFR dropped to 4.4% on the 14^th^ day since the declared beginning of the outbreak on February 19, 2020. Later, between 15 March 2020 and 12 April 2020, the CFR stabilized between 5.2% and 7.8% with a mean of 6.7%. See figure 3(c). With the wider spread of COVID-19 across the country, the CFR increased to 7.86% on 23 March 2020. However, the CFR declined and plateaued around 6.2% between 1 and 15 April 2020. The relatively high CFR could correspond to a significant level of under-reporting of the infected cases and an overwhelmed health system. Given the wide distribution of the time from confirmation-to-death of COVID-19, we also explore the delay-adjusted CFR with 5- and 10-day delay period, as examples. The dynamics of the delay-adjusted CFR with 10-day delay suggests that the CFR has been gradually reducing in Iran from 17.97% on 16 March 2020 to 8.20% on 12 April 2020.

When underreporting of infected cases is overlooked, the estimated effective reproduction number began from 5.67 (95% CI 5.48-4.86) on 1 March 2020 and reduced to 0.70 (95% CI 0.69-0.71) on 15 April 2020 suggesting the outbreak peak has already occurred on 8 April 2020 when goes below 1, about 50 days from the confirmation of the first case. The outbreak is also likely to continue until the end of 2020. See figure 4. The estimates of the effective reproduction number were consistently larger during the early stage of the outbreak when underreporting is overlooked compared to when underreporting is taken into account. However, the estimated effective reproduction numbers converged as the number of infected cases approached the peak. The convergence of the estimates can be partly explained by the fact that the effective reproduction number is more dependent on the rate of change in the infected and recovered cases rather than their absolute numbers. Results also suggest that the impact of control measures on the effective reproduction number is significantly overestimated when under-reporting is taken into account.

**Figure 4:**
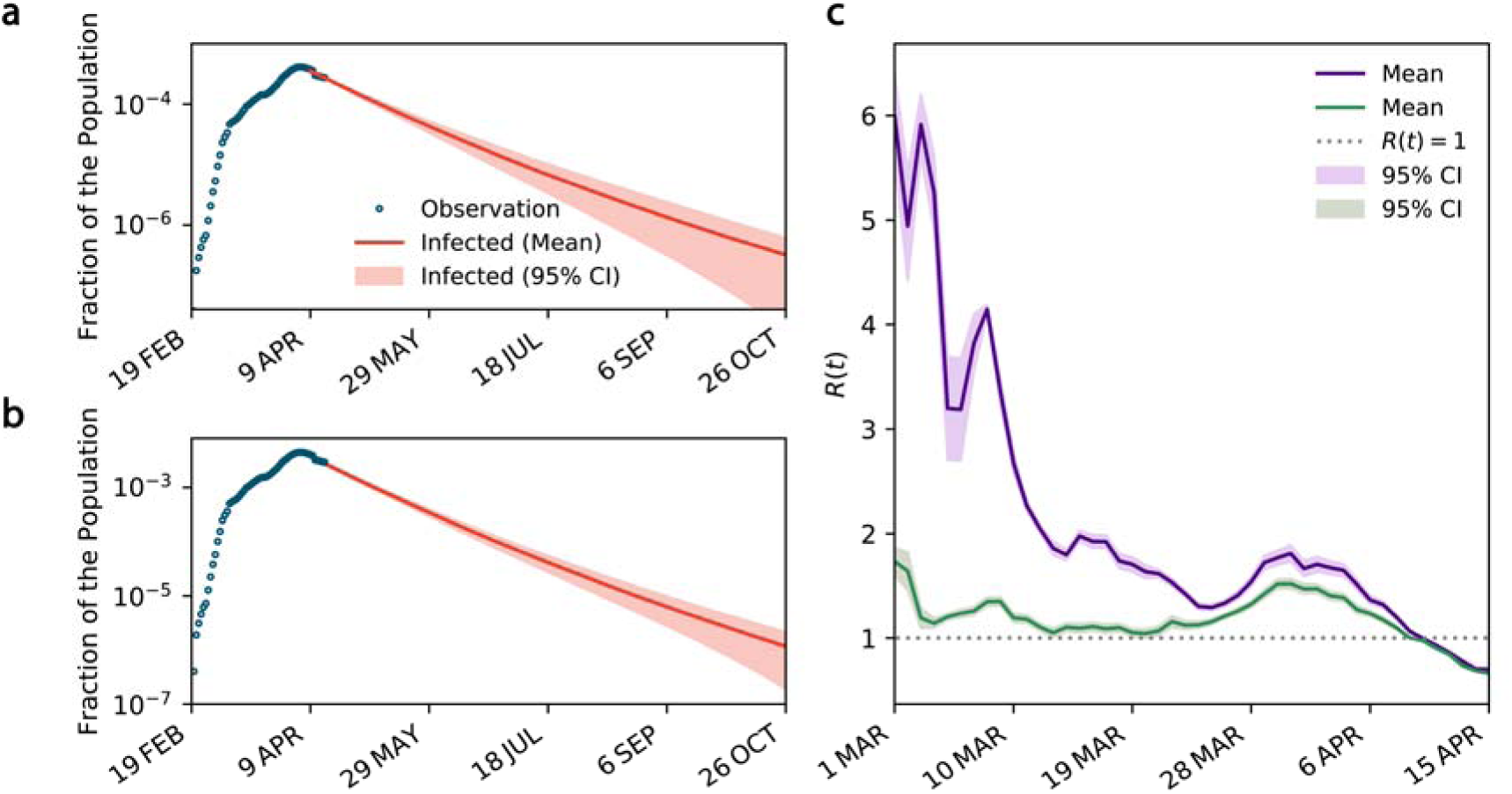
The impact of under-reporting on the estimated dynamics of transmission. (a) and (b) The projected curves for the number of infected cases when underreporting is not considered and when it is taken into account, respectively. (c) Temporal variations of *R*(*t*) when underreporting is overlooked (purple curve) and when it is taken into account (green curve).

With the gradual reduction of the effective reproduction number to below one and the increasing pressure on an already fragile economy because of the implemented control measures, the government is seeking an exit strategy and is considering easing some of the restrictions. Here, we conduct a scenario analysis to understand how three different scenarios could change the projected outlook of the outbreak in Iran: i) maintaining the same level of control measures as of 12 April 2020, ii) intensifying the measures to increase physical distancing represented by a 20% reduction in the reproduction number, and iii) partially lifting the restrictions to ease physical distancing represented by a 20% increase in the reproduction number. To estimate the number of ICU beds needed, we assume 5% of the confirmed cases require intensive care.^20^ We found that in all scenarios the projection of patients requiring ICU admission exceeds the original ICU capacity. Note that no official information is available on the expanded ICU capacity. As of 12 April 2020, both projected curves start above the ICU capacity. Easing the restrictions can quickly push the peak to a level that is five times higher than the scenario where the current level of control measures is maintained and puts additional pressure on the health system. Results clearly suggest that with further restrictions (scenario ii), the projected curve quickly goes back under the ICU capacity while it takes more than 100 days for the curve to go back under the ICU capacity in scenario i and iii.

**Figure 5:**
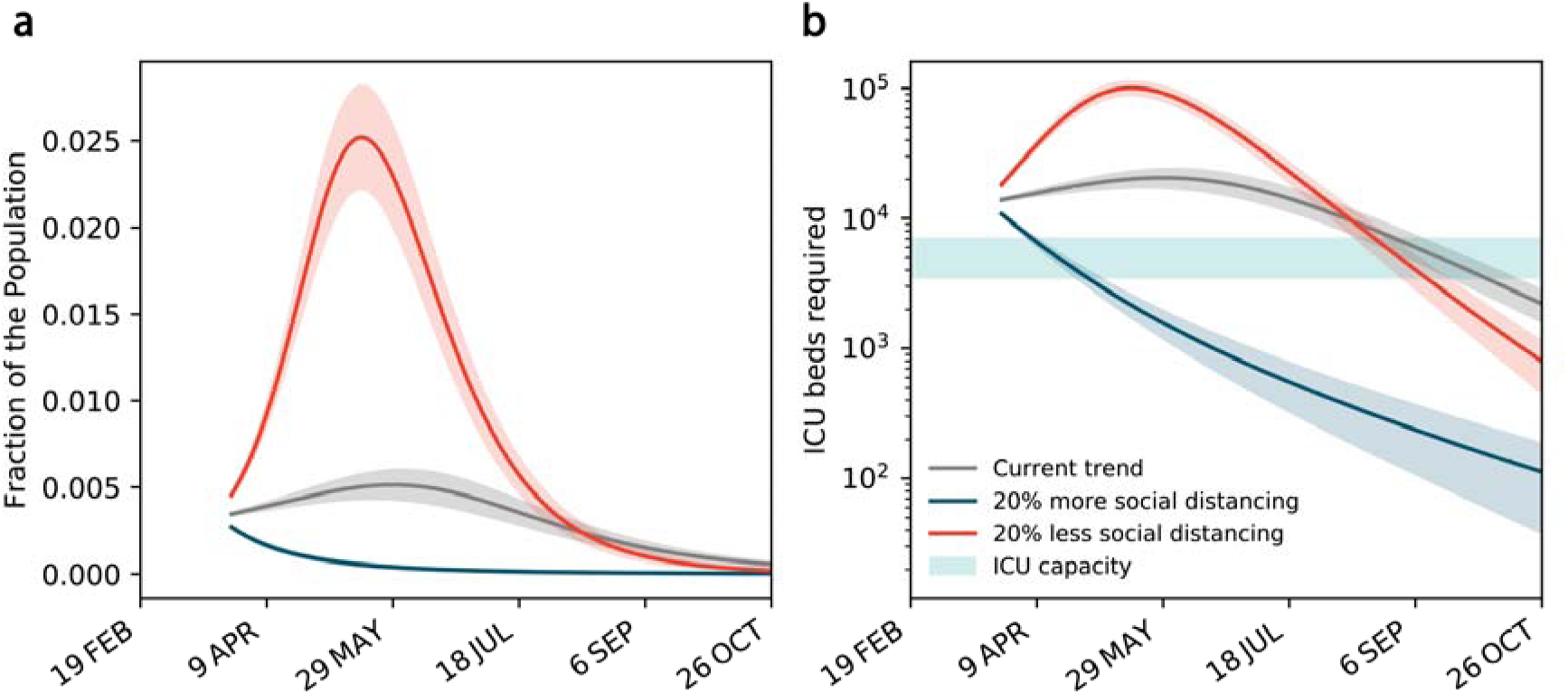
Scenario analysis for maintaining, reinforcing or easing the restrictions. (a) The projected curves for the total number of confirmed and suspected cases under three different scenarios: maintaining the same level of control measures as of 12 April 2020, reinforcing the control measures to increase physical distancing by a 20% increase in the reproduction number, and partial lifting the restrictions to ease physical distancing by a 20% decrease in the reproduction number. (b) The projected number of ICU beds needed under the three scenarios. The original ICU capacity is assumed between 3500 to 7000 beds.^7^

## DISCUSSION

In this study, we used a mathematical epidemic model to provide the first estimates of the changing transmission of SARS-CoV-2 infection in Iran when underreporting of cases are considered. We used official data and adjusted our estimates for underreporting based on delay-adjusted CFR.

We used a variation of the SEIR model combined with an estimate of the percentage of symptomatic cases using delay-adjusted CFR based on the distribution of the time from confirmation-to-death^19^. In contrast to previous models of the epidemic^21^, we did not assume any prior information on the distribution of the effective reproduction number. This combined with the use of a series of distributions allowed us to take into account more appropriately the uncertainty and the random variation in both input data and model outcomes.

The outbreak of COVID-19 in Iran is claimed to have been started in Qom province with the first case officially reported on 19 February 2020. As shown earlier in figure 1(a), according to the official data, it only took two days, for four more provinces, including Tehran, Markazi, Mazandaran, and Gilan (Guilan) to report their first cases. The first non-pharmaceutical intervention took place on 22 February 2020 with the closure of schools and universities in Qom followed by the capital city of Tehran after a day. With the school closures in the capital and yet low level of social awareness about the potential risks of COVID-19, a surge in the number of holiday trips from Tehran to northern provinces of Mazandaran and Gilan was observed. Soon after, with no restriction on the inter-city travels the number of identified cases in Gilan and Mazandaran grew rapidly, and the infection spread throughout the rest of the country.

The second major non-pharmaceutical intervention occurred on 28 February 2020 with a wide campaign to disinfect public spaces and closure of all schools and universities across all provinces. On 5 March 2020, the government announced an increase in the COVID-19 testing capacity. On 7 March 2020, sporadic closure or reduction of working hours in government offices and banks across the country was reported. On 14 March 2020, two often crowded religious shrines in Qom and Mashhad were closed to visitors. The increased physical distancing as the result of interventions and increase in public awareness of the crisis through major official and unofficial information campaigns, especially on social media, gradually showed its impact by slowing down the speed of the spread as shown earlier in figure 1(b). The estimated effective reproduction number increased from 1.14 on 4 March 2020 to 1.35 on 8 March 2020, perhaps due to the increase in case ascertainment and delay in early interventions to show their impact. The effective reproduction number decreased consistently to 1.04 on 20 March 2020.

On 17 March 2020, only a few days before the Persian new year (Nowruz, 20 March 2020), in the absence of strict travel restrictions, millions of Iranians began making road trips to various destinations across the country despite the warnings from the government and many hotels, restaurants and the general hospitality industry’s refrainment to provide services to any traveller. This is believed to have increased the speed of the spread of COVID-19 in Iran, increasing the effective reproduction number to 1.52 on 30 March 2020.

On 22 March 2020, government announced more restrictive non-pharmaceutical interventions leading to closure of all non-essential businesses for at least two weeks, followed by further intensified interventions on March 28, 2020 including restrictions on entry and exit to affected provinces and cities, closure of parks, pools and all recreational places, and a ban on public gatherings including sport, cultural, and religious events. These restrictions pushed the effective reproduction number further down to 1.0 on 9 April 2020 and 0.69 on 15 April 2020.

Iran has had one of the highest case fatality rates (CFR) among the affected countries in the world. While the CFR is known to vary significantly between countries due to various reasons including testing frequency and population age distribution^18^, the CFR is still higher in Iran despite having a younger population with the median age of 30.8 years old compared to China with 4% CFR and the median population age of 37.4 and South Korea with 2% CFR and the median population age of 41.8.^18^ Perhaps a comparable country in the Middle East region with similar population characteristics (median age of 30.9) is Turkey. The CFR in Turkey as of 6 April 2020 was 2%, three times lower than that of Iran.^8^ While the evidence is indirect, it suggests that the official number of cases reported by Iran may has been significantly under-reported, possibly due to relatively low case-ascertainment and under-reporting of identified cases.^18^

The continuous reduction in the delay-adjusted CFR shows a different trend compared to the CFR with no consideration of the time from confirmation-to-death. The continuous reduction could be explained by the improving case identification practice in Iran over time with various initiatives including a National Coronavirus Helpline (the “4030” service), established in late February 2020 to self-report symptoms and identify suspected cases.

Our results confirmed the significant impact of underreporting in describing the story of the COVID-19 outbreak in Iran, especially in the early stages. We showed how overlooking underreporting can drastically affect estimation of *R(t)* and overestimate the impact of control measures.

Our results also showed the reduction in effective reproduction number, a measure of infection transmission, during this period. This decrease was most likely due to the increased physical distancing as the result of multiple non-pharmaceutical public health interventions, including school closures, ban on public gatherings, travel restrictions, full/partial closure of non-essential businesses, as well as major awareness campaigns over social media. Based on the latest trends, while the first peak of COVID-19 in Iran occurred on 5 April 2020, the post-peak period may continue to the end of 2020. However, these projections assume the continuation of the current level of control measures and the absence of effective therapeutic treatment or vaccination programs. Hence, they can be subject to important shifts depending, in particular, on the public’s willingness to continue and the government’s success in implementing social distancing measures or easing the restrictions.

Our model provides tangible evidence of the association between the different non-pharmaceutical interventions and their impact on the course of the outbreak. The results showed how the acceptance and hence effectiveness of the interventions endorsed by the Iranian government aimed at “flattening the curve” depended in part on the public’s level of awareness of the principles behind governmental policies and their trust in the government and its control measures. For example, closure of schools and businesses in the capital city of Tehran near the time of the Persian new year was followed by a surge of holiday trips to Gilan and Mazandaran and a subsequent increase in the number of cases in these provinces and in other parts of the country. This observation shows how interventions may be associated with unintended and at times counterproductive consequences. These negative consequences can be prevented when there is open and credible communications by competent officials and mutual trust between public and government. Moreover, intervention measures need to be developed, implemented and enforced as a whole, for example by strict reinforcement of travel restrictions in conjunction with school and workplace closings.

There is no substitute for high quality data – complete, accurate, and timely – as a basis for public policy. However, in the absence of such data, modelling of the type presented here can help provide reasonable estimates as well as realistic bounds of their uncertainty. The range of uncertainty can be viewed as the margin of error in the model’s predictions of the number of cases, ICU admissions or deaths. Modeling can also illustrate different scenarios -- pessimistic vs. optimistic vs. realistic -- of how the epidemic may evolve in relation to current and future public health measures and the possible compliance of the public over time.

In conclusion, using a stochastic model of the SARS-CoV-2 epidemic in Iran, we assessed the dynamic of the epidemic in relation to public health measures to increase social distancing. We took into account both the inherent uncertainty in the data and the possible impact of under-reporting of true cases due to low case ascertainment and reporting. In the absence of consistently reliable data, the modelling approach as presented here can help generated reasonable estimates of key public health metrics such as the number of cases and case fatality ratio. In turn, these metrics and scenarios can help serve the dual purpose of informing public policy and the public and fostering discussions and improvements of epidemic modelling.

## Data Availability

All data used in the modelling are available upon request.

